# Performance of Contemporary Large Language Models on the National Prosthodontic Resident Examination: A Multi-Year Comparative Study

**DOI:** 10.64898/2026.10.19.26365937

**Authors:** Yo-wei Chen, Vathanai Bhunyanaphakul, Pitchaporn Kosago, Matthew Finkelman, Damian J. Lee

**Author notes:** Corresponding author: Dr. Yo-wei Chen DDS, DMD, MSc, PhD, FACP, Department of Prosthodontics, Tufts University School of Dental Medicine, Boston, Massachusetts, USA, 1 Kneeland St. Suite 1248, Boston, MA, 02111. Yo-wei Chen and Vathanai Bhunyanaphakul contributed equally to this work and share joint first authorship.

## Abstract

**Purpose:** To compare the performance of three large language models (LLMs)—ChatGPT-o1, Grok 3, and Gemini 2.0 Pro—on the American College of Prosthodontists’ National Prosthodontic Resident Examination (NPRE).

**Materials and Methods:** Official NPRE examinations from years 2015-2024 were retrieved from the American College of Prosthodontists website and used to prompt each LLM. Paid access tiers and advanced reasoning were enabled. Responses from LLMs were recorded and compared against the official answer keys for accuracy, variability across years, and consistency of responses. Duplicate questions were retained for the primary analysis and removed in a sensitivity analysis. Accuracy (percent correct) was compared using generalized estimating equations binary logistic regression (primary); Cochran’s Q with Bonferroni-adjusted McNemar tests (sensitivity). Year-to-year variability (SD of annual accuracy) was compared using weighted Levene tests. Consistency across duplicate items was quantified as the percentage of perfect matches and compared using Cochran’s Q with Bonferroni-adjusted McNemar tests.

**Results:** Across 1,487 questions, overall accuracy differed among models (global P < 0.001). ChatGPT-o1 achieved 78.4% correct, Grok 3 76.4%, and Gemini 2.0 Pro 73.2%; ChatGPT-o1 and Grok 3’s accuracy significantly exceeded Gemini 2.0 Pro (both P ≤ 0.016), with no significant difference between ChatGPT-o1 and Grok 3 (P = 0.099). After removing duplicates (n = 1,200), results were similar: ChatGPT-o1, Grok 3, and Gemini 2.0 Pro were 77.1%, 76.6%, and 71.5% correct, respectively; ChatGPT-o1 and Grok 3’s accuracy was significantly higher than Gemini 2.0 Pro (both P < 0.001). Variability across years did not differ significantly (P ≥ 0.097). Grok 3 showed significantly higher perfect-match consistency (79.2%) than ChatGPT-o1 (69.4%) and Gemini 2.0 Pro (64.5%) (P ≤ 0.010).

**Conclusions:** Contemporary LLMs showed 70–80% accuracy on postgraduate prosthodontic multiple-choice content. While promising for educational support, residual error and inconsistency warrant human oversight and transparent methods reporting before curricular adoption.

## Introduction

Large language models (LLMs) have gained profound popularity in recent years due to advances in artificial intelligence (AI), especially in the generative AI aspect (1–4). Technology can generate human language and perform reasoning tasks based on prompts (5). These models are capable of performing automation processes, supporting users in various tasks, and decision-making assistance for improving workflows in widespread sectors (1). In education, the abilities to comprehend sophisticated inquiries, and report evidence and supportive argument responses are invaluable for both students and instructors in providing decision making and problem-solving (6, 7).

Various companies have developed and released their own LLMs. ChatGPT (OpenAI, San Francisco, CA, USA), Gemini (Google, Mountain View, CA, USA), and Grok (xAI, San Francisco, CA, USA) are three commonly used LLMs. ChatGPT, developed by OpenAI, is a general-purpose LLM trained on extensive internet-scale data, widely applied in education, customer service, and automation tasks. Gemini, developed by Google DeepMind, is a multimodal model designed to process and integrate text, images, and code, with a strong focus on research, reasoning, and productivity applications. Grok, developed relatively recently by xAI, is notable for its close integration with the social platform X, emphasizing rapid information retrieval and conversational engagement, with a stated commitment to promoting open expression.

Recent studies reported the performance of LLMs on national medical examinations, such as the United States Medical Licensing Exam (USMLE)(3) and Polish Medical Final Examination(4). They found that the LLMs can display comprehensible reasoning and valid clinical knowledge, as well as explanatory responses (3). Although the performance was less than the human-exam takers, they reported the performance as “near or at the passing score of the tests” (3, 4). These studies showed the possibilities of integrating LLMs into dental education. One study subjected LLMs to Oral and Maxillofacial Surgery Board Exams and found that the overall score across models was 69.69%, doubting the performance to be sufficient for clinical and educational use. Further training and refinement were recommended (8).

In Advanced Education in Prosthodontic Programs (AEPP) in the United States, prosthodontics residents take the National Prosthodontic Resident Examination (NPRE) with a series of multiple-choice questions to attest their capability and assessed knowledge in prosthodontic specialty. Passing of the NPRE is often used by the program directors to evaluate the resident’s overall performance level and the program’s effectiveness as an outcome measure. Additionally, the format and make-up of the questions are similar to the American Board of Prosthodontics section A examination. The American College of Prosthodontists (ACP) provides a set of questions to residents and their test results every year, followed by official answers and references for them to review the materials. Each examination comprises 150 postgraduate-level prosthodontic questions on different topics, such as removable prosthodontics, fixed prosthodontics, implant dentistry, and maxillofacial prosthetics, among others. Reviewing these topics and questions is crucial for residents to prepare for their board examination and inform dental practice. Several studies (2, 6, 9) have demonstrated that the accuracy of LLMs in Prosthodontics ranged from 44% to 64% and was inconsistent over time, limiting their reliability as independent educational resources. Although individual models may exhibit strengths based on question type, subtopic, or repeatability, their performance often fluctuates, and repeated exposure to correct information does not consistently enhance subsequent outputs. These findings underscore both the promise of AI models as adjunctive educational tools and the need for further refinement, before they can serve as dependable sources of Prosthodontic knowledge. A recent study reported a score of 73.7% performance in 2024 NPRE exam using ChatGPT 4(10). However, to the authors’ knowledge, there is no study comparing the performance of different LLMs in postgraduate level prosthodontics examinations in multiple years.

Therefore, the aim of this study was to comparatively evaluate the performance of different LLMs, namely ChatGPT-o1, Grok 3, and Gemini 2.0 Pro, to provide answers from postgraduate prosthodontics questions according to the NPRE. The performance in terms of correctness of the given answers, variability of performance across question years, and the consistency of each LLM to provide the same response to questions that were duplicated across years were analyzed. The null hypothesis was that the three models would show equal performance in responding to NPRE questions, regarding correctness, variability, and consistency.

## Materials and Methods

To evaluate the accuracy, variability, and consistency of the performance of the LLMs, the National Prosthodontic Resident Examinations (NPRE) and their answer sheets from year 2015 – 2024 were retrieved from the ACP website (11), which were accessible to members of the College. All questions were provided with an option list, and test takers were asked to answer using multiple-choice responses. In cases where a question was duplicated (i.e., when the same question appeared in multiple years), the questions were entered into each LLM once for every year it appeared.

Three LLMs were selected for this study including ChatGPT-o1 (paid access tier), Grok 3 (paid subscription), and Gemini 2.0 Pro (paid access tier), which were widely used and popular generative AI models. The paid versions of those LLMs were used in this study with the advanced reasoning model features enabled. The prompt given to the LLMs was to provide answers for each examination. To standardize behavior across LLMs, a simple prompt was provided “Please take the exam, provide answers for all 150 questions.” All prompting occurred between March 1st and May 27, 2025. The responses provided from each year’s examination were recorded and compared with the answers given by the ACP. Examinations were provided to the LLMs starting from the most recent year, 2024, descending to 2015. Each examination year was run in a fresh conversation to minimize context carryover. All questions and answers were stored in an electronic spreadsheet (Excel Professional Plus 2024; Microsoft Corp.). All answers provided by the LLMs were compared with the original answers.

The performance of different LLMs was observed in terms of accuracy, variability, and consistency. Accuracy was evaluated by the overall percentage of correct answers to the entire set of questions from all years, while variability was defined as the spread of the performance across the examination years. Consistency was observed by determining whether each LLM provided the same answer each time a duplicate question was asked.

For performance accuracy, the percentages of correct answers were compared between LLMs. In the primary analysis, this was done without removing the duplicate questions; generalized estimating equations (GEE) binary logistic regression was conducted to account for the presence of correlated data. In addition, a sensitivity (secondary) analysis was conducted in which the LLMs’ percent accuracies were compared when removing the duplicate questions, to evaluate whether conclusions were different with and without the presence of duplicates. In the sensitivity analysis, the first answer provided by the LLM was kept for the analysis; Cochran’s Q test was used for the global test, and McNemar’s test was used in post-hoc comparisons. For both the primary and the sensitivity analysis, the significance level was set at 0.05 for the global test, whereas the Bonferroni correction was used in post-hoc comparisons.

Regarding the comparison of LLMs in terms of variability of performance across the years, each LLM’s percentage of correct answers was calculated for each year. The standard deviation of each of these percentages was then computed for each LLM. In the primary analysis (in which data from duplicate questions were included), the significance test comparing the LLMs was performed via the weighted Levene’s test, using inverse error variances as weights and 10,000 bootstrap samples to estimate the null distribution, accounting for correlated data from the duplicates. The sensitivity analysis (removing the duplicates) also used the weighted Levene’s test, using inverse error variances as weights; however, bootstrap samples were not needed to estimate the null distribution due to the removal of duplicates.

Lastly, the consistency in providing answers was evaluated using the duplicate questions. Answers from the same questions listed in different examination years were collected and compared for the consistency of the answers given. The LLMs were compared in terms of the percentage of perfect matching, i.e., the percentage of duplicate questions for which the LLM always provided the same answer whenever the duplicate question was asked. Cochran’s Q test was used for the global test in conjunction with a significance level of 0.05; McNemar’s test with the Bonferroni correction was used for post-hoc comparisons. No sensitivity analysis was conducted for this aim because it could only be addressed using duplicate data. Statistical analyses were conducted using SAS 9.4 (SAS Institute Inc., Cary, NC, USA) and SPSS 30 (IBM Corp., Armonk, NY, USA).

## Results

### Comparison of overall percent correctness across LLMs

A total of 1487 questions spanning ten years, including duplicate items, were evaluated in this study. Thirteen questions in examination year 2015 were excluded, as the questions had been removed from grading as indicated in the answer sheet provided by the ACP. A statistically significant difference was observed in the global test comparing the LLMs (P = 0.001) (Table 1). Overall, ChatGPT-o1 achieved the highest accuracy at 78.4%, followed by Grok 3 at 76.4%, and Gemini 2.0 Pro at 73.2%. In post-hoc comparisons, both ChatGPT-o1 (P < 0.001) and Grok 3 (P = 0.016) answered a significantly greater percentage of questions correctly than Gemini 2.0 Pro. However, the difference between ChatGPT-o1 and Grok 3 was not significant (P = 0.099). The sensitivity analysis of accuracy after removal of duplicated questions was consistent with the primary accuracy findings (Table 2). A total of 1200 questions were analyzed. ChatGPT-o1 demonstrated a correctness of 77.1%, followed by Grok 3 at 76.6% and Gemini 2.0 Pro at 71.5%. While there was no significant difference between ChatGPT-o1 and Grok 3 (P = 0.752), both performed significantly better than Gemini 2.0 Pro (P < 0.001). **Figure 1** illustrates a comparison of primary and sensitivity analysis among LLMs.

**Figure 1:**
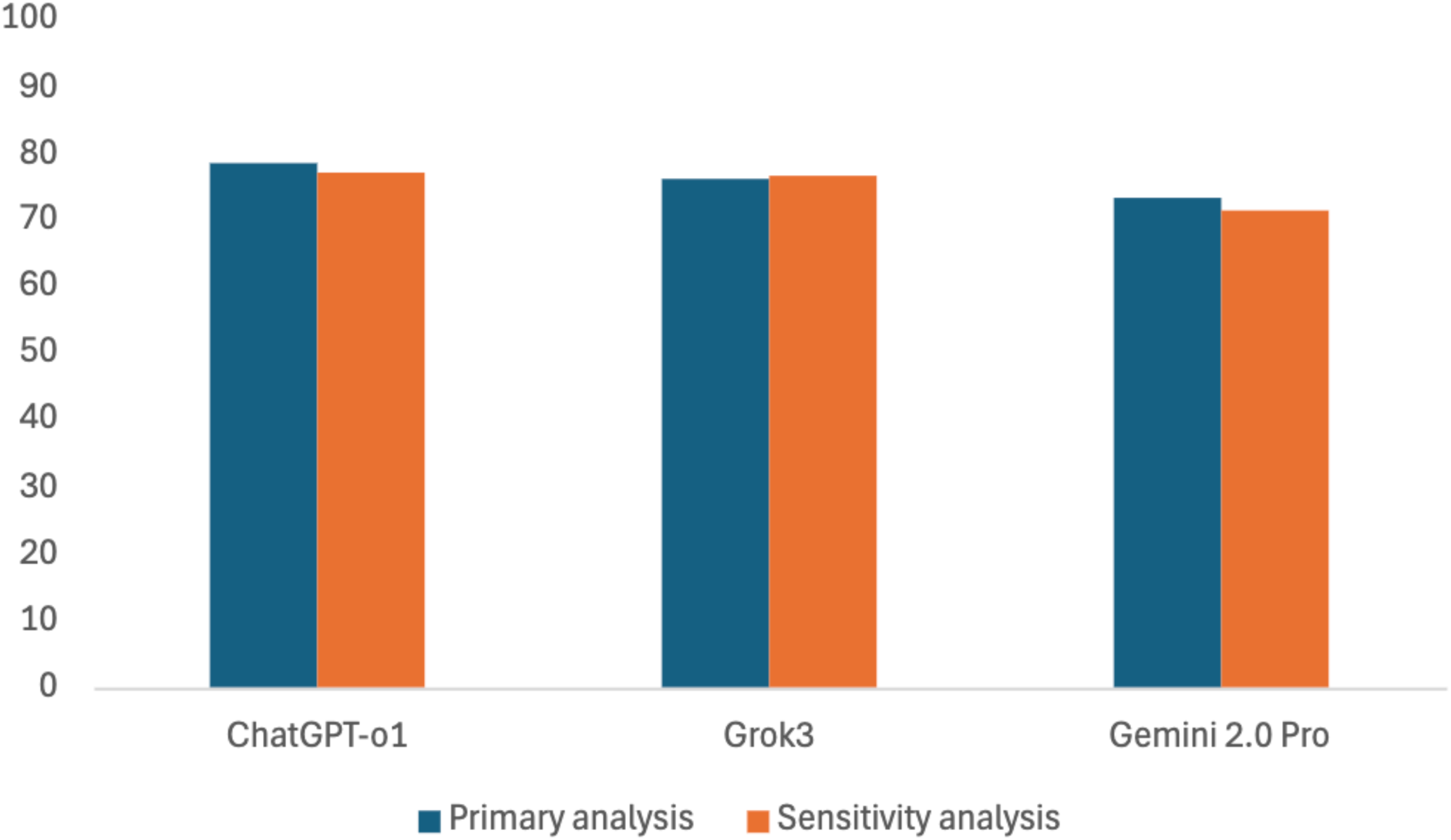
Overall accuracy of large language models across all NPRE questions (primary and sensitivity analysis)

**Table 1:** Performance of LLMs in terms of overall percent correctness without removing the duplicates (primary analysis)

| Large language<br>model (LLM) | Correct answer |  | Incorrect answer |  | p-value |
| --- | --- | --- | --- | --- | --- |
|  | Count | Percentage | Count | Percentage |  |
| **ChatGPT-o1** | 1166 | 78.4<sup>a</sup> | 321 | 21.6 | 0.001* |
| **Grok 3** | 1136 | 76.4<sup>a</sup> | 351 | 23.6 |  |
| **Gemini 2.0 Pro** | 1088 | 73.2<sup>b</sup> | 399 | 26.8 |  |

\*p-value represents result from GEE binary logistic regression global test
Each superscript denotes a subset of groups that did not significantly differ from each other

**Table 2:**
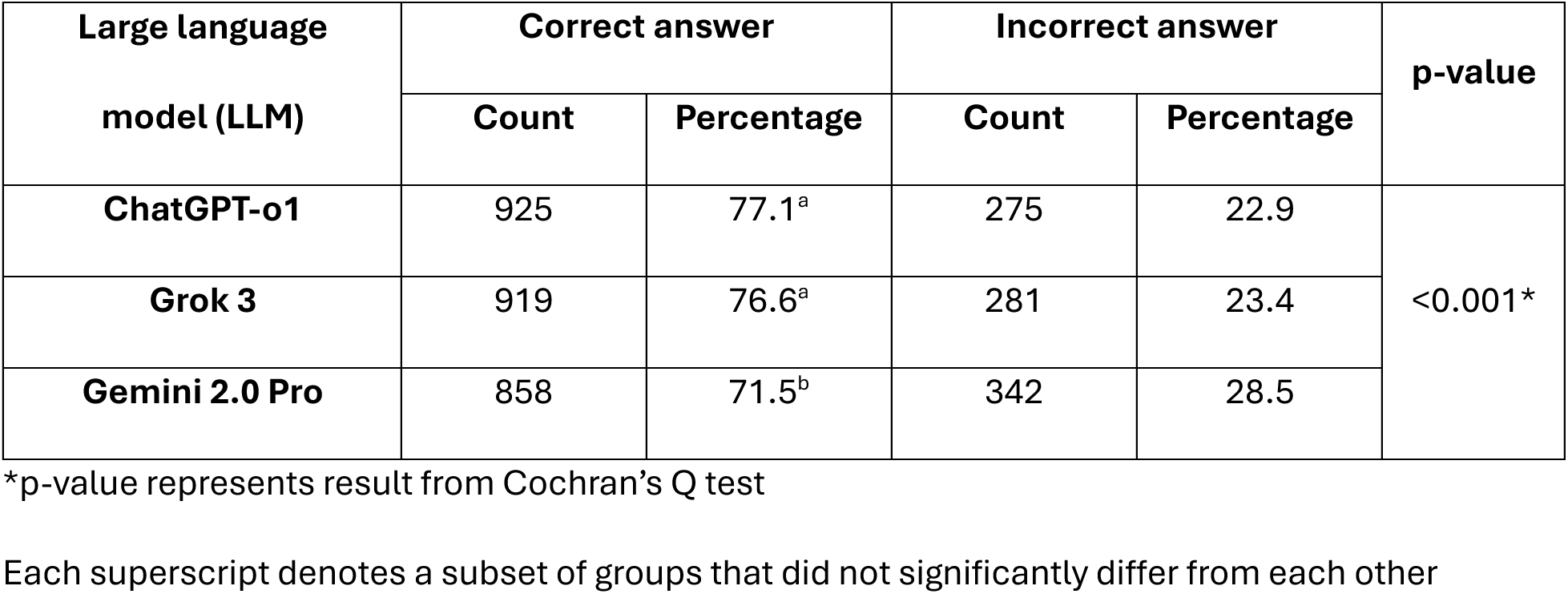
Performance of LLMs in terms of overall percent correctness with duplicates removed (sensitivity analysis)

| Large language<br>model (LLM) | Correct answer |  | Incorrect answer |  | p-value |
| --- | --- | --- | --- | --- | --- |
|  | Count | Percentage | Count | Percentage |  |
| **ChatGPT-o1** | 925 | 77.1<sup>a</sup> | 275 | 22.9 | <0.001* |
| **Grok 3** | 919 | 76.6<sup>a</sup> | 281 | 23.4 |  |
| **Gemini 2.0 Pro** | 858 | 71.5<sup>b</sup> | 342 | 28.5 |  |

\*p-value represents result from Cochran's Q test
Each superscript denotes a subset of groups that did not significantly differ from each other

### Comparison of variability of between year performance among LLMs

Comparison of LLMs in terms of variability of performance across years is presented in **Table 3** and **Figure 2**. In the primary analysis (including duplicates), standard deviations for ChatGPT-o1, Grok 3, and Gemini 2.0 Pro’s percentages of correct answers were 4.4%, 7.7%, and 9.1%, respectively. No statistically significant difference in variability across years among the LLMs was observed (P = 0.097). In the sensitivity analysis, in which duplicates were removed, the standard deviations were 6.5%, 8.1%, and 10.4% for ChatGPT-o1, Grok 3, and Gemini 2.0 Pro, respectively. As in the primary analysis, the sensitivity analysis (shown in **Table 4**) removing duplicates showed no significant difference between the LLMs (P = 0.331).

**Figure 2:**
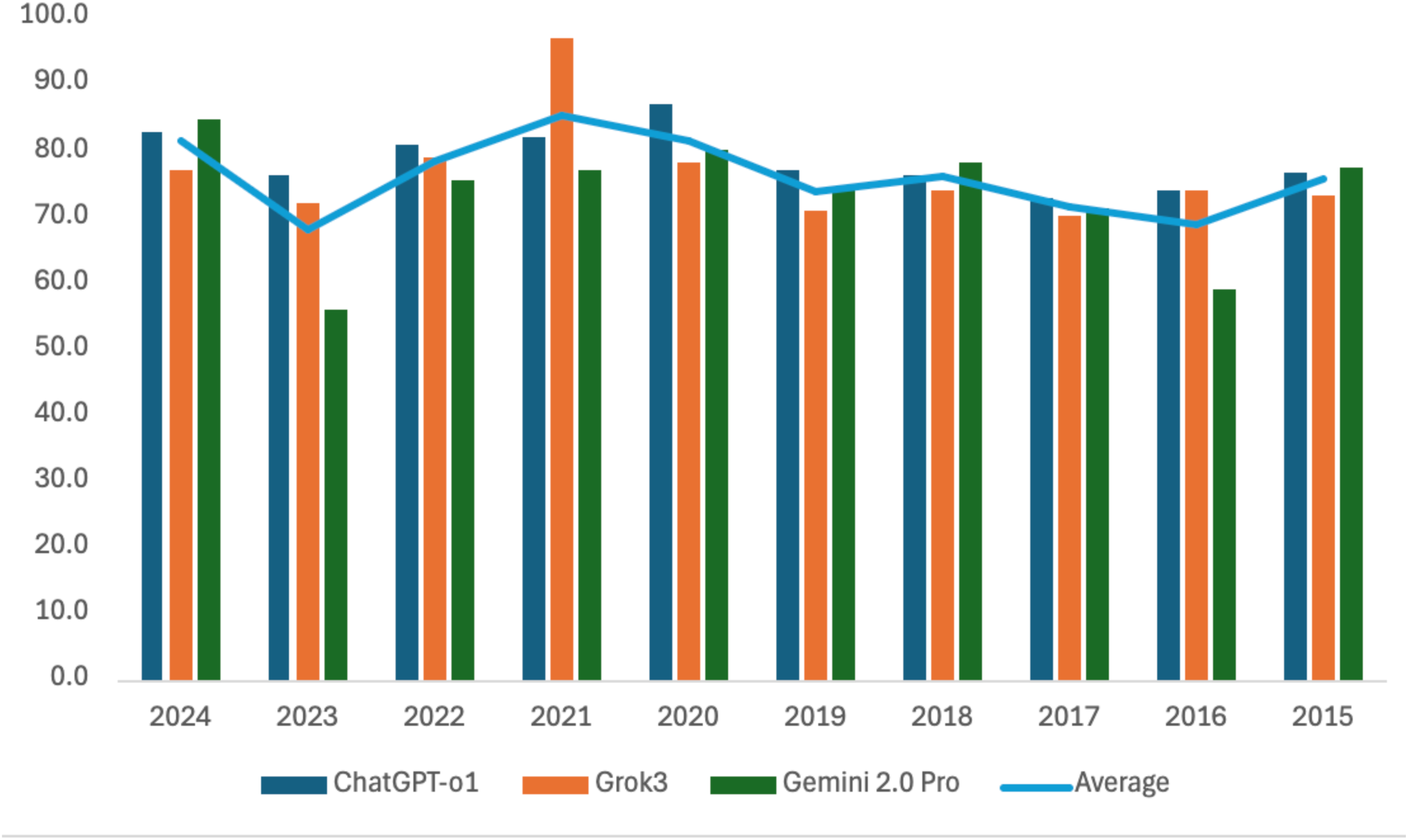
Variability of accuracy of large language models across examination years

**Table 3:**
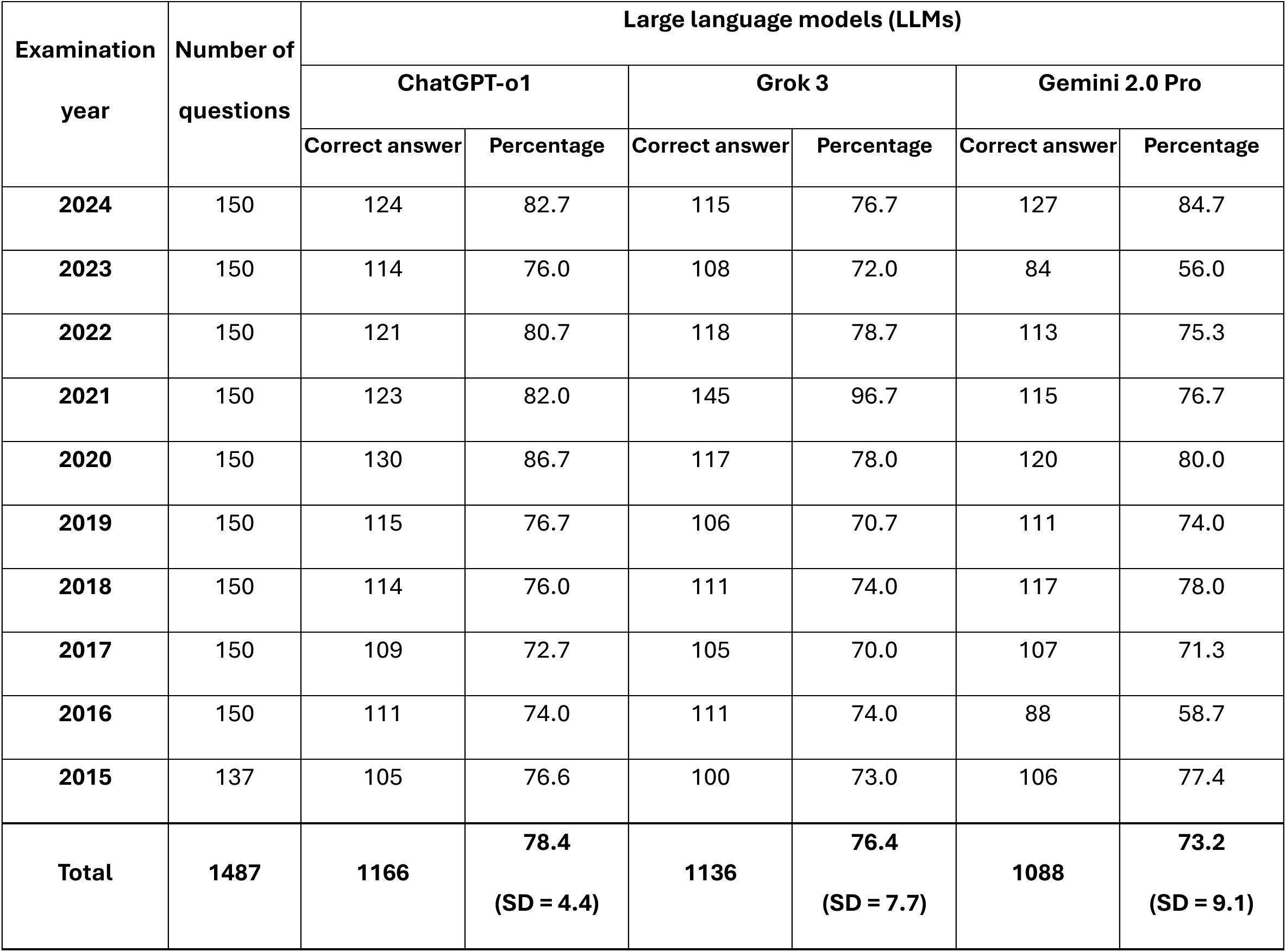
Variability of LLM performance across years without removing the duplicates (primary analysis)

**Table 4:** Variability of LLM performance across years with duplicates removed (sensitivity analysis)

| Examination<br>year | Number of<br>questions | Large language models (LLMs) |  |  |  |  |  |
| --- | --- | --- | --- | --- | --- | --- | --- |
|  |  | ChatGPT-o1 |  | Grok 3 |  | Gemini 2.0 Pro |  |
|  |  | Correct<br>answer | Percentage | Correct<br>answer | Percentage | Correct<br>answer | Percentage |
| 2024 | 150 | 124 | 82.7 | 115 | 76.7 | 127 | 84.7 |
| 2023 | 141 | 106 | 75.2 | 101 | 71.6 | 77 | 54.6 |
| 2022 | 138 | 111 | 80.4 | 107 | 77.5 | 102 | 73.9 |
| 2021 | 129 | 108 | 83.7 | 124 | 96.1 | 98 | 76.0 |
| 2020 | 131 | 111 | 84.7 | 102 | 77.9 | 103 | 78.6 |
| 2019 | 98 | 64 | 65.3 | 66 | 67.3 | 71 | 72.4 |
| 2018 | 99 | 70 | 70.7 | 79 | 79.8 | 75 | 75.8 |
| 2017 | 107 | 77 | 72.0 | 73 | 68.2 | 72 | 67.3 |
| 2016 | 89 | 64 | 71.9 | 66 | 74.2 | 46 | 51.7 |
| 2015 | 118 | 90 | 76.3 | 86 | 72.9 | 87 | 73.7 |
| Total | 1200 | 925 | 77.1<br>(SD = 6.5) | 919 | 76.6<br>(SD = 8.1) | 858 | 71.5<br>(SD = 10.4) |

### Comparison of LLMs in terms of their consistency of answers for duplicated questions

A total of 245 questions and their duplicates were used in this analysis. Grok 3 achieved the highest percentage of perfect matching across duplicates at 79.2%, compared with ChatGPT-o1 (69.4%) and Gemini 2.0 Pro (64.5%). There was a statistically significant difference in the global test comparing the LLMs (P < 0.001). In post-hoc comparisons, Grok 3’s percentage was significantly higher than both ChatGPT-o1 (P = 0.010) and Gemini 2.0 Pro (P < 0.001); however, the difference between ChatGPT-o1 and Gemini 2.0 Pro was not significant (P = 0.230).

## Discussion

This study demonstrated the performance of three LLMs in their paid versions when applied to the context of prosthodontics. Many aspects of their performances, including accuracy, variability, and consistency, were used to determine their potential to be used as a study guide and provide accurate and useful information at the residency level. This level of questions challenged the LLMs with advanced, sophisticated, and research-based information that would need extensive searching and a critical thinking strategy to provide an answer. Accurate, stable, and consistent responses are imperative for LLMs to be effective (12).

Statistical analysis revealed significant differences among the LLMs. ChatGPT-o1 and Grok 3 achieved statistically significantly higher accuracy than Gemini 2.0 Pro. Although residents’ scoring performance is not publicly available and thus could not be compared with human averages, all LLMs exceeded 70% accuracy on the examinations. This is consistent with a previous systematic review indicating that LLMs could show different levels of performance in different studies(13). In addition, this finding is aligned with the results reported by Shirani (2) that focused on the fixed prosthodontics questions compared between ChatGPT-4o, DeepSeek R1, and Gemini 2.0 Pro. ChatGPT-4o demonstrated significantly better performance and remained stable throughout the study period. However, performance differed depending on the type of question; LLMs performed better in multiple-choice questions than in short-answer type. In contrast, our findings differed from those of a previous study (6), which reported that ChatGPT-3.5 performed similarly to Gemini, with no significant difference in accuracy across dental subtopics. This discrepancy may be attributed to the use of the free version of ChatGPT-3.5 in the previous study, as different versions exhibit varying performance characteristics (14). Furthermore, topic-specific accuracy differences may have contributed to the variation (6, 14, 15). For instance, Eraslan et al. (6) noted the highest accuracy in implantology and the lowest in removable partial dentures, possibly due to the latter’s reliance on older, textbook-based knowledge that may not be well-represented in current LLM training data. These findings highlight the need for further investigation into LLM performance across diverse dental subdomains.

Variability across examination years provides insight into the reproducibility of LLM performance. Lower variability suggests more stable performance when LLMs are exposed to postgraduate prosthodontic content. In the present study, between-model differences in variability were not statistically significant. Variability is a shared limitation across LLMs, which are influenced by training data, system updates, and the probabilistic nature of generative AI outputs (16). Particularly in postgraduate education, where reliability is critical, such fluctuations underscore the importance of continuous benchmarking of models. Until stability of performance is demonstrated, along with the accuracy of the provided responses, LLMs should be positioned as supplementary tools rather than stand-alone study resources (12).

Consistency of the LLMs to provide the same answer to a duplicated question is crucial in many aspects. Especially in the health care setting, consistency of responses impacts patient safety, education, and decision making of the healthcare provider. On the other hand, confusion and misinformation may occur with inconsistency of the results (17). This present study showed that different LLMs provided various degrees of consistency in terms of perfect matching, approximately 64 – 80%. This level of consistency may be considered unsatisfactory and may be attributed to the probabilistic nature of LLMs (16).

Previous research (18) showed a different degree of consistency achieved by ChatGPT4, with 85.4% in answering endodontic questions, while the current study reported the consistency of ChatGPT-o1 at 69.4%. The prior study used the yes/no question type and asked LLMs to provide 60 answers per question at different times of day. In their study, consistency was identified by the proportion of matching responses. In contrast, the present study quantified consistency as the presence or absence of perfect agreement across all answers to the same question, which is crucial in higher education, where fluctuations in responses are undesirable. Question type may also impact the consistency of responses (19). Although the prior study (19) stated that yes/no and multiple-choice questions demonstrated similar performance, more complex question types introduced greater challenges for the LLMs. The different features and logic of LLMs may contribute to varying levels of consistency when responding to different question types. With different models and methodologies used, direct comparison or similarity of the results may be difficult. In an educational context, these inconsistencies create potential risks for students who may rely on AI outputs when studying for examinations or making clinical judgments (12).

Most LLMs do not fully disclose their training methods (20), though they are believed to use a mix of textbooks, peer-reviewed articles, internet resources, and other materials to acquire scientific knowledge. The filtering process for these sources remains unclear. Newer versions of LLMs generally outperform older ones; however, even the highest accuracy observed in this study (78.4%) indicates a risk of error. In patient care, such an error rate is unacceptable without human oversight. Therefore, current LLMs should serve as a complement to, rather than a replacement for, expert judgment in treatment planning.

The current study has several limitations. First, only three major LLMs were tested, and they were not released at the same time. With the continuous development of LLMs, newer models tend to outperform older ones. Second, only multiple-choice questions were used. Compared to open-ended questions, multiple-choice items may be easier for LLMs to answer correctly. Since the focus of this study was not on evaluating the reasoning behind the answers, future studies should assess the validity of the models’ explanations or include open-ended questions. Image-based questions were also excluded here; incorporating different question types would provide a more comprehensive evaluation of LLM performance across diverse tasks. Third, the prompts used in this study were not consistent across models due to differences in how they generate responses. For example, some LLMs required clarification that the task was for research purposes; otherwise, they would not provide answers due to user guidelines. Fourth, this study tested only the paid versions of LLMs, which may not be accessible to all users. Limitations on usage and capacity are usually applied to free versions. Future research comparing paid and free versions would provide valuable insights into their relative performance for a broader user base. Therefore, the results of this study should be interpreted with caution.

## Conclusion

Modern LLMs have demonstrated a performance threshold that enables credible, though not flawless, support for postgraduate prosthodontic education. ChatGPT-o1, Grok 3, and Gemini 2.0 Pro showed promising accuracy in answering questions in NPRE examinations. Their integration into resident training could possibly streamline formative assessment and personalized remediation, provided robust safeguards remain in place to mitigate residual errors and opacity in reasoning. Continuous monitoring, explainability auditing, and domain-specific fine-tuning will be critical steps toward the responsible adoption of generative AI in prosthodontic practice and education.

## Data Availability

All data produced in the present study are available upon reasonable request to the authors

The NPRE questions and answer keys are the property of the American College of Prosthodontists and are available to ACP members; they cannot be redistributed by the authors. The LLM responses and scoring data are available from the corresponding author upon reasonable request.

## Acknowledgements

The authors would like to thank Tyler Karp for analytical assistance.

## Ethics statement

This study did not involve human participants, human data, or human tissue; institutional review board approval was therefore not required. The American College of Prosthodontists granted permission to use the National Prosthodontic Resident Examination (NPRE) materials for this research.

